# ROCCA study protocol and interim analysis on safety of Sputnik V vaccine (Gam-COVID-Vac) in the Republic of San Marino: an observational study using active surveillance

**DOI:** 10.1101/2021.05.03.21256509

**Authors:** Marco Montalti, Giorgia Soldà, Zeno Di Valerio, Aurelia Salussolia, Jacopo Lenzi, Marcello Forcellini, Edoardo Barvas, Susanna Guttman, Rossella Messina, Elisabetta Poluzzi, Emanuel Raschi, Rossano Riccardi, Maria Pia Fantini, Giusy La Fauci, Davide Gori, for the San Marino Republic COVID ROCCA Group

## Abstract

**OBJECTIVE:** To assess the safety of the Sputnik V (Gam-COVID-Vac) COVID-19 vaccine through participant-based active surveillance from the Republic of San Marino vaccination campaign.

**DESIGN AND SETTING:** This is a nation-wide cohort study in the Republic of San Marino to monitor any Adverse Events Following Immunisation (AEFI) with the Sputnik V.

**PARTICIPANTS:** Adults aged 18-89 years who had at least one dose of Sputnik V administered and who responded or accessed to an e-questionnaire sent via email, QR-code or were live/phone interviewed about the 7 days after the first vaccine dose (n=2,558) and 7 days after the second dose (n=1,288). Exclusion criteria were inability to understand nor to answer the questionnaire properly.

**MAIN OUTCOME MEASURES:** Rates of overall AEFI short-term and long-term (3 months). Secondary outcomes focus on subgroups of the population presenting specific comorbidities. Being this an interim analysis, long-term data (3 months) is still being collected, especially for vulnerable populations, including subjects with comorbidities and the youngest age groups.

**RESULT:** The median age of participants was 68 years. 56% were females. After the first dose, vaccine recipients described both local and systemic reactions in 16.4% of cases, 25.8% reported systemic reactions only, and 10.2% reported local symptoms only. After the second dose, both local and systemic reactions were reported in 31.9% of cases, 18.5% reported systemic reactions only, and 16.1% reported local symptoms only. Main symptoms were local pain (24.8% for first dose and 43.8% for the second), asthenia (23.8% and 31.9%), headache (18.5% and 21.0%), and joint pain (16.5% and 21.9%). In the population over 60, recipients having reported AEFI after the first dose could be a predictor of AEFI recurrence after the second dose (p<0.001). 81.8% of those reporting second-dose AEFI, reported AEFI after the first dose, while amongst those not having reported any AEFI after the first dose, 18.2% reported AEFI after the second dose.

**CONCLUSION:** The ROCCA interim analysis confirmed a good tolerability profile in the over 60 years age group after both doses regarding short-term solicited AEFI to Sputnik V (Gam-COVID-Vac).

## Introduction

One year after the onset of the COVID-19 pandemic, deaths approach 3 millions worldwide [1]. The Republic of San Marino was among the first countries to be struck by the still ongoing COVID-19 pandemic, and also one of the countries with the highest death toll if compared with its population [2].

Faced with the dire consequences of the pandemic, this country has decided to employ all the available resources in order to reach the population coverage needed to grant herd immunity. The Sputnik V vaccine was included in its vaccination campaign [3].

The Sputnik V vaccine, the first to be registered among anti-COVID vaccines, is a heterologous recombinant adenovirus vaccine with two different types of adenovirus vectors (rAd26 and rAd5) for the first and second dose respectively, developed by the Gamaleya National Center of Epidemiology and Microbiology in Moscow [4].

It was registered in August 2020 by the Russian Ministry of Health as Gam-COVID-Vac [5] and starting from December 2020 mass distribution in several countries took place including Argentina, Belarus, Hungary, Serbia and the United Arab Emirates. The list of countries where Sputnik V has been registered has since then expanded, reaching the current number of 61, including Russia [6].

On February 23, the Republic of San Marino received its first supply of Sputnik V, launching the national vaccination campaign on the 25th of the same month [7] starting from health professionals and most vulnerable citizens. At 55 days from the beginning 46.1% (n=15,621) of the population has been vaccinated for the first dose [8] with the goal of a rapid vaccination campaign for the entire population.

In the process of planning a national vaccination campaign, the acquisition of solid evidence on vaccine efficacy and safety is of the utmost importance.

The only available phase 3 trial on Sputnik V showed a 91.6% (95%CI; 85.6-95.2) efficacy with no significant differences assessed in the age strata, and 100% (95%CI; 94.4-100.0) efficacy against moderate or severe COVID-19 [9].

Evidence deriving from clinical trials preceding approval of vaccines is essential but not sufficient. In fact, it needs to be complemented in a real world setting by an immediate and continuous stream of information gathered alongside the campaign progression.

Vaccine pharmacovigilance (i.e., post-marketing safety monitoring of vaccines) fills this gap, being defined as the science and activities relating to the detection, assessment, understanding and communication of Adverse Events Following Immunisation (AEFI) and other vaccine- or immunisation-related issues, and to the prevention of untoward effects of the vaccine or immunisation [10].

The aim of this study is to present our ROCCA^1^ research protocol for active vaccine surveillance (i.e., solicited reporting) and to show the preliminary data on prevalence and characteristics of AEFI to the Sputnik V vaccine among the population of the Republic of San Marino.

## Methods

### Study design and Participants

This is a nation-wide cohort study to assess safety of Sputnik V anti-COVID vaccine in the Republic of San Marino. The vaccination process includes one dose of rAd26-S (0.5 ml) administered intramuscularly on day 0 and one dose of rAd5-S (0.5 ml) administered intramuscularly on day 21. Both the vaccination hubs set up for the campaign by the Social Security Institute (SSI) are involved for participants recruitment.

All the people vaccinated with Sputnik V are actively recruited by physicians immediately after the administration of the vaccine. Eligibility criteria have been defined as: age over 18, having had at least one dose of anti-COVID vaccine administered, being covered by the SSI health insurance. Exclusion criteria have been defined as: not being able to understand nor to answer the questionnaires properly. No randomisation or special selection was carried out. All participants provided informed consent to being included in the database for study participation.

### Outcomes

The primary outcome measure has been defined as safety of the Sputnik V (Gam-COVID-Vac) COVID-19 vaccine, measured as the number of participants reporting AEFI, in the first week after the first dose, in the first week after the second dose, and - in the long term - within 3 months after the first dose. This enables the collection of patient-reported safety data in near real time and long term and generates incident rate of AEFI.

Secondary outcomes focus on detecting AEFI on subgroups of the population presenting specific comorbidities (eg. diabetes, neurological diseases).

### Procedure and Questionnaire

The standardized e-questionnaires are administered actively to the participants to collect information about potential AEFI following vaccine injection. The questionnaires have been generated using Google Form, and can be filled in autonomously or with the help of a member of the research team.

Vaccine recipients are asked to fill in the questionnaires at fixed time intervals, namely: 1 week (Q1), 1 month (Q2), 3 months after the first vaccine dose (Q3) (Figure S1).

Q1 investigates demographic information (*patient code*; *age; weight; height; gender; profession*), anamnestic data (*pregnancy; diseases; therapy; recent vaccinations; allergies; previous COVID-19*), date of the first injection, vaccine brand, the potential AEFI occurring in the week after the first dose, and the severity/impact of the symptoms (including need for medical assistance and hospitalization). Q1 includes 7 sections for a total of 25 questions with closed mandatory answers, while Q2 and Q3 contain 2 sections for a total of 6 questions. In order to link the questionnaires one to another, the patient code is asked also in Q2 and in Q3. Q2 investigates the potential AEFI occurring in the week after the second dose. Q3 investigates further potential AEFI occuring in the long-term period after the vaccination and it will be administered starting from June 2021.

Answers to questions concerning relevant variables were made mandatory in order to complete the questionnaire to minimize missing data.

The list of AEFI and most of the questions were adapted from vaccine surveillance studies conducted by European Medicines Agency. The events indicated are in line with the typical vaccine adverse reactions already identified by the relevant Regulatory Agencies, as well as for the latency of possible occurrence.

The clinical features of any AEFI, their frequency and intensity will be specified on the questionnaires using the CTCAE scale (Common Terminology Criteria for Adverse Events) version 5.0.

An informed consent form was attached to the e-questionnaire, and each participant consented to participate in the survey after reading the information sheet.

### Data collection

Time frame for data collection lasts from the 4th of March 2021 to the end of the national vaccination campaign. The collection timeframe for data included in this preliminary analysis was from 4th of March to the 8th of April, 2021. Data collection is carried out with three different approaches: face-to-face interviews at vaccination sites, telephone interviews, and online access to the e-questionnaires which will be guaranteed either by QR-code scanning, or through a link sent via email. Sensitive information collected will hence be stored anonymously in order to guarantee participants privacy.

### Medical history assessment

The modest population size of San Marino Republic allows for all study participants to be connected via their SSI code to their clinical records allowing linkage with anamnestic data and drug use, besides allowing the verification of the accesses to the national health service following the vaccination.

### Statistical analysis

Numerical variables were summarised as mean ± standard deviation; categorical variables were summarised as frequencies and percentages. Frequency distributions were depicted with the aid of bar charts and frequency polygons. McNemar’s test was used to evaluate differences in the occurrence of adverse reactions after the first and the second shot of the vaccine. Differences between distinct groups of individuals were assessed using the two-sample test of proportions for large samples (i.e., asymptotically normal).

In this interim analysis, having recruited a total of 1946 individuals aged 60–89 years from a population of 8799 (source: https://www.statistica.sm), our sample size was sufficient to estimate a proportion of adverse events equal to 1% ± 0.4% up to 50% ±2% with a confidence level of 95% [11]. All analyses were carried out using Stata software, version 15 (StataCorp, 2017, *Stata Statistical Software: Release 15*, College Station, Texas, USA: StataCorp LP). The significance level was set at 5%.

## Results

### Population characteristics

We recruited 2,558 participants who received Sputnik V (Gam-COVID-Vac) between February 25, 2021, and April 8, 2021. Among these, 56% (n=1,424) were females and 19.4%(n=496) were obese (Body Mass Index ≥ 30). Mean age was 66 years, median age was 68 years, and the most represented age group was 60-69 years (45.4%, n=576) (Table 1, Figure 1).

**Table 1.** Sociodemographic characteristics of the study sample, overall and by age group - Republic of San Marino (2021). Values are counts (percentages) or mean ± standard deviation.

| Characteristic | All<br>(n = 2558) | Age group |  |  |  |  |  |
| --- | --- | --- | --- | --- | --- | --- | --- |
|  |  | 18–39 y<br>(n = 183) | 40–49 y<br>(n = 177) | 50–59 y<br>(n = 252) | 60–69 y<br>(n = 786) | 70–79 y<br>(n = 799) | 80–89 y<br>(n = 361) |
| Female | 1424 (55.7) | 103<br>(56.3) | 111<br>(62.7) | 176<br>(69.8) | 427<br>(54.3) | 427<br>(53.4) | 180 (49.9) |
| Age, y | 65.6 ± 14.3 | 30.4 ± 5.4 | 44.5 ± 2.9 | 54.7 ± 2.8 | 65.2 ± 2.7 | 74.8 ± 2.9 | 82.1 ± 1.6 |
| BMI, kg/m<sup>2</sup> | 26.5 ± 4.8 | 23.8 ± 4.6 | 25.4 ± 5.5 | 25.9 ± 5.8 | 27.1 ± 4.6 | 26.9 ± 4.5 | 26.8 ± 3.8 |
| BMI category,<br>kg/m<sup>2</sup> |  |  |  |  |  |  |  |
| <18.5 | 41 (1.6) | 12 (6.6) | 3 (1.7) | 3 (1.2) | 2 (0.3) | 15 (1.9) | 6 (1.7) |
| 18.5–<25 | 997 (39.0) | 112<br>(61.2) | 96 (54.2) | 133<br>(52.8) | 272<br>(34.6) | 276<br>(34.5) | 108 (29.9) |
| 25–<30 | 1017 (39.8) | 43 (23.5) | 55 (31.1) | 73 (29.0) | 334<br>(42.5) | 335<br>(41.9) | 177 (49.0) |
| 30–<35 | 374 (14.6) | 11 (6.0) | 10 (5.6) | 28 (11.1) | 133<br>(16.9) | 134<br>(16.8) | 58 (16.1) |
| 35–<40 | 89 (3.5) | 3 (1.6) | 7 (4.0) | 6 (2.4) | 37 (4.7) | 28 (3.5) | 8 (2.2) |
| ≥40 | 33 (1.3) | 2 (1.1) | 5 (2.8) | 8 (3.2) | 8 (1.0) | 9 (1.1) | 1 (0.3) |
| Unspecified | 7 (0.3) | 0 (0.0) | 1 (0.6) | 1 (0.4) | 0 (0.0) | 2 (0.3) | 3 (0.8) |
| Healthcare worker | 289 (11.3) | 79 (43.2) | 72 (40.7) | 98 (38.9) | 39 (5.0) | 1 (0.1) | 0 (0.0) |
| Non-physician<br>profession* | 157 (6.1) | 51 (27.9) | 39 (22.0) | 53 (21.0) | 14 (1.8) | 0 (0.0) | 0 (0.0) |
| Clerk | 98 (3.8) | 22 (12.0) | 28 (15.8) | 34 (13.5) | 14 (1.8) | 0 (0.0) | 0 (0.0) |
| Physician | 23 (0.9) | 3 (1.6) | 3 (1.7) | 6 (2.4) | 10 (1.3) | 1 (0.1) | 0 (0.0) |
| Pharmacist | 7 (0.3) | 2 (1.1) | 2 (1.1) | 3 (1.2) | 0 (0.0) | 0 (0.0) | 0 (0.0) |
| Volunteer | 4 (0.2) | 1 (0.5) | 0 (0.0) | 2 (0.8) | 1 (0.1) | 0 (0.0) | 0 (0.0) |

\* Nurse, midwife, physical therapist, podiatrist, speech-language pathologist, orthoptist, audiologist, dental hygienist, dietician, etc.
BMI, body mass index.

**Figure 1.**
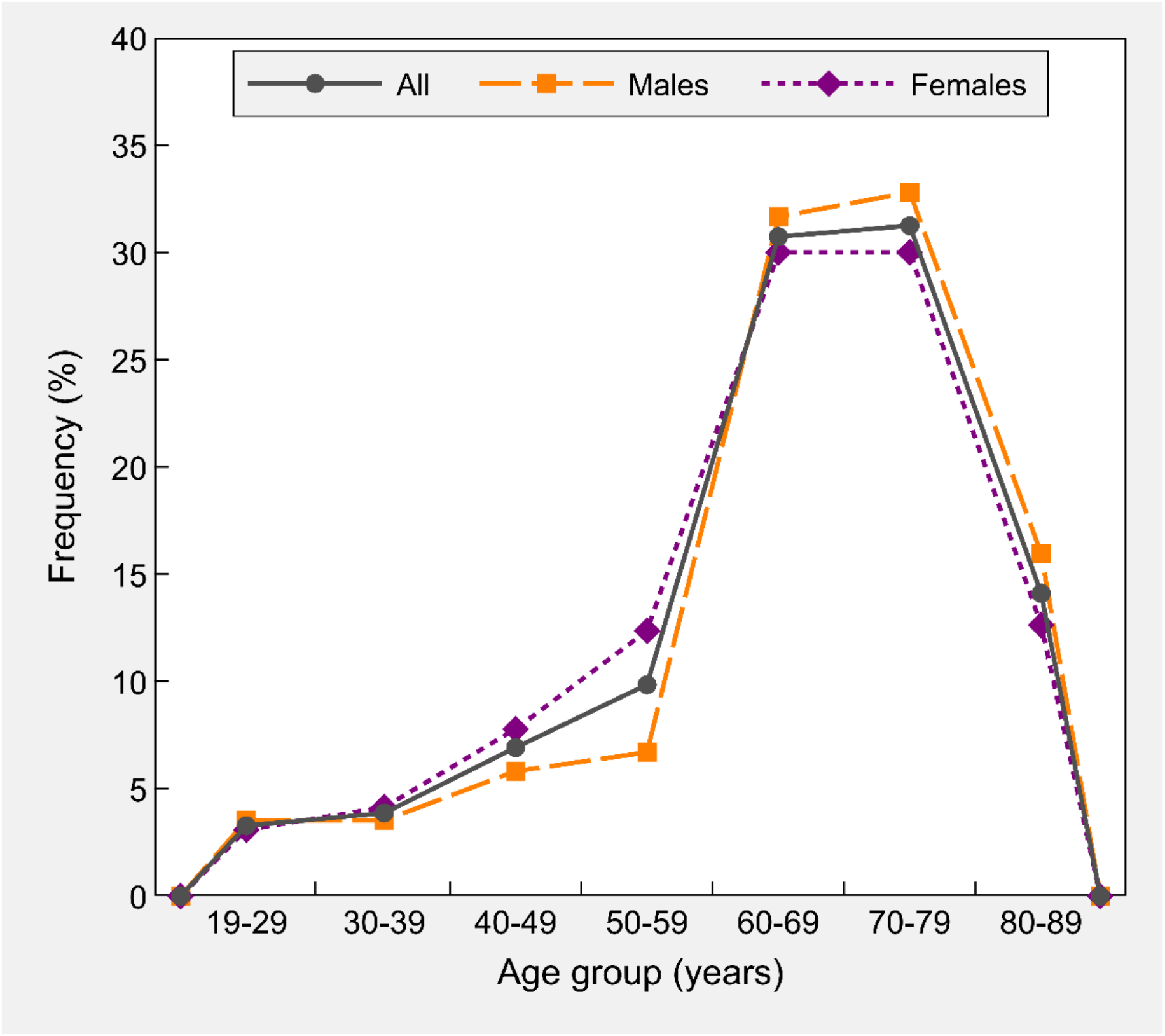
Percentage age distribution of the study sample (n = 2558), overall and by sex - Republic of San Marino (2021).

Among these 2,558 participants, 75.1% (n=1,920) had at least one underlying medical condition. Hypertension was the most frequent coexisting condition (46.4%, n=1,188), followed by cardiovascular diseases (32.1%, n=822). A previous infection with SARS-CoV-2 was reported in 3.6% (n=93) of the participants. Participants suffering from at least one allergy were 24.5% (n=627), most frequently to drugs (12%, n=307) (Table 2).

**Table 2.** Baseline clinical characteristics of the study sample, overall and by age group - Republic of San Marino (2021). Values are counts (percentages).

| Characteristic | All<br>(n = 2558) | Age group |  |  |  |  |  |
| --- | --- | --- | --- | --- | --- | --- | --- |
|  |  | 19–39 y<br>(n = 183) | 40–49 y<br>(n = 177) | 50–59 y<br>(n = 252) | 60–69 y<br>(n = 786) | 70–79 y<br>(n = 799) | 80–89 y<br>(n = 361) |
| Underlying medical conditions | 1920 (75.1) | 44 (24.0) | 50 (28.2) | 147 (58.3) | 591 (75.2) | 736 (92.1) | 352 (97.5) |
| Hypertension | 1188 (46.4) | 3 (1.6) | 13 (7.3) | 58 (23.0) | 347 (44.1) | 507 (63.5) | 260 (72.0) |
| Cardiovascular diseases | 822 (32.1) | 2 (1.1) | 5 (2.8) | 34 (13.5) | 221 (28.1) | 345 (43.2) | 215 (59.6) |
| Osteoarticular diseases | 388 (15.2) | 1 (0.5) | 3 (1.7) | 11 (4.4) | 65 (8.3) | 185 (23.2) | 123 (34.1) |
| Diabetes mellitus | 297 (11.6) | 6 (3.3) | 5 (2.8) | 16 (6.3) | 79 (10.1) | 126 (15.8) | 65 (18.0) |
| Mental disorders | 181 (7.1) | 2 (1.1) | 5 (2.8) | 14 (5.6) | 34 (4.3) | 79 (9.9) | 47 (13.0) |
| Respiratory diseases | 171 (6.7) | 9 (4.9) | 3 (1.7) | 12 (4.8) | 30 (3.8) | 73 (9.1) | 44 (12.2) |
| Neurological diseases | 159 (6.2) | 8 (4.4) | 7 (4.0) | 11 (4.4) | 29 (3.7) | 57 (7.1) | 47 (13.0) |
| Malignant tumours | 132 (5.2) | 0 (0.0) | 1 (0.6) | 11 (4.4) | 24 (3.1) | 57 (7.1) | 39 (10.8) |
| Nephropathy | 77 (3.0) | 0 (0.0) | 0 (0.0) | 5 (2.0) | 8 (1.0) | 27 (3.4) | 37 (10.2) |
| Immunosuppression | 65 (2.5) | 3 (1.6) | 2 (1.1) | 4 (1.6) | 26 (3.3) | 22 (2.8) | 8 (2.2) |
| Liver diseases | 52 (2.0) | 1 (0.5) | 1 (0.6) | 3 (1.2) | 9 (1.1) | 25 (3.1) | 13 (3.6) |
| Other* | 779 (30.5) | 23 (12.6) | 20 (11.3) | 63 (25.0) | 203 (25.8) | 315 (39.4) | 155 (42.9) |
| Allergies | 627 (24.5) | 64 (35.0) | 59 (33.3) | 90 (35.7) | 167 (21.2) | 180 (22.5) | 67 (18.6) |
| Drugs | 307 (12.0) | 15 (8.2) | 19 (10.7) | 34 (13.5) | 95 (12.1) | 106 (13.3) | 38 (10.5) |
| Rhinitis | 209 (8.2) | 47 (25.7) | 30 (16.9) | 39 (15.5) | 40 (5.1) | 39 (4.9) | 14 (3.9) |
| Contact dermatitis | 85 (3.3) | 7 (3.8) | 12 (6.8) | 12 (4.8) | 28 (3.6) | 18 (2.3) | 8 (2.2) |
| Food | 59 (2.3) | 6 (3.3) | 8 (4.5) | 7 (2.8) | 16 (2.0) | 14 (1.8) | 8 (2.2) |
| Asthma | 52 (2.0) | 13 (7.1) | 7 (4.0) | 8 (3.2) | 12 (1.5) | 9 (1.1) | 3 (0.8) |
| Insect sting | 27 (1.1) | 1 (0.5) | 4 (2.3) | 4 (1.6) | 9 (1.1) | 5 (0.6) | 4 (1.1) |
| Other | 16 (0.6) | 2 (1.1) | 3 (1.7) | 4 (1.6) | 2 (0.3) | 4 (0.5) | 1 (0.3) |
| Ongoing drug therapies | 1847 (72.2) | 50 (27.3) | 46 (26.0) | 134 (53.2) | 569 (72.4) | 710 (88.9) | 338 (93.6) |
| Other vaccines in the two weeks before | 5 (0.2) | 0 (0.0) | 0 (0.0) | 0 (0.0) | 0 (0.0) | 1 (0.1) | 4 (1.1) |
| Painkillers/anti-inflammatory drugs before vaccination | 88 (3.4) | 8 (4.4) | 4 (2.3) | 6 (2.4) | 17 (2.2) | 25 (3.1) | 28 (7.8) |
| Previous infection with SARS-CoV-2 |  |  |  |  |  |  |  |
| No | 2465 (96.4) | 166 (90.7) | 165 (93.2) | 231 (91.7) | 761 (96.8) | 785 (98.2) | 357 (98.9) |
| Asymptomatic | 14 (0.5) | 3 (1.6) | 2 (1.1) | 2 (0.8) | 5 (0.6) | 2 (0.3) | 0 (0.0) |
| Mild symptoms | 46 (1.8) | 11 (6.0) | 7 (4.0) | 7 (2.8) | 10 (1.3) | 7 (0.9) | 4 (1.1) |
| Moderate/severe symptoms | 22 (0.9) | 2 (1.1) | 2 (1.1) | 12 (4.8) | 4 (0.5) | 2 (0.3) | 0 (0.0) |
| Severe symptoms with hospitalisation | 11 (0.4) | 1 (0.5) | 1 (0.6) | 0 (0.0) | 6 (0.8) | 3 (0.4) | 0 (0.0) |

\* Thyroid disorders, prostate disorders, gastroesophageal reflux disease, glaucoma. SARS-CoV-2, severe acute respiratory syndrome coronavirus 2.

### First dose AEFI

Vaccine recipients described both local and systemic reactions in 16.4% of cases; besides, 25.8% reported systemic reactions, and 10.2% reported local symptoms only. Most frequent local reactions were pain (24.8%), nodules (3.7%), warmth (2.2%), swelling (1.9%). The most frequent systemic reported symptoms were asthenia (23.8%), headache (18.5%), joint pain (16.5%), chills (16.5%), muscle pain (16%), fever (11.9%), and malaise (11.8%), (Figure 2, 3). For other AEFI, not included in the provided list, see the Table S1.

**Figure 2.**
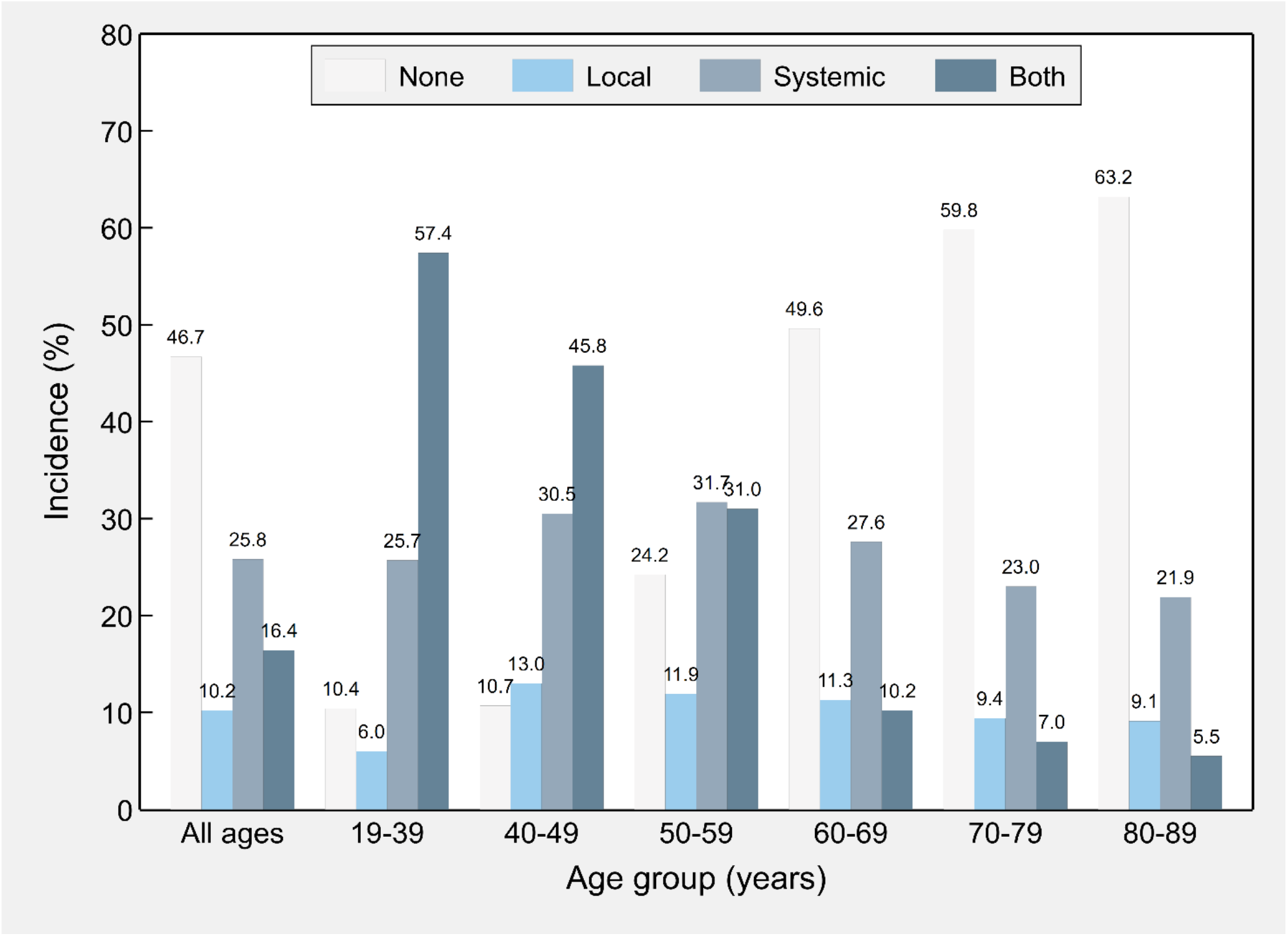
Incidence of local and systemic adverse events following immunisation with the first shot of Sputnik V (n = 2558), overall and by age group - Republic of San Marino (2021). *Notes:* Percentages of each age group do not add up to 100 because unspecified symptoms were not included in the chart (all: 0.8%; 19–39 y: 0.5%; 40–49 y: 0.0%; 50–59 y: 1.2%; 60–69 y: 1.3%; 70–79 y: 0.8%; 80–89 y: 0.3%).

**Figure 3.**
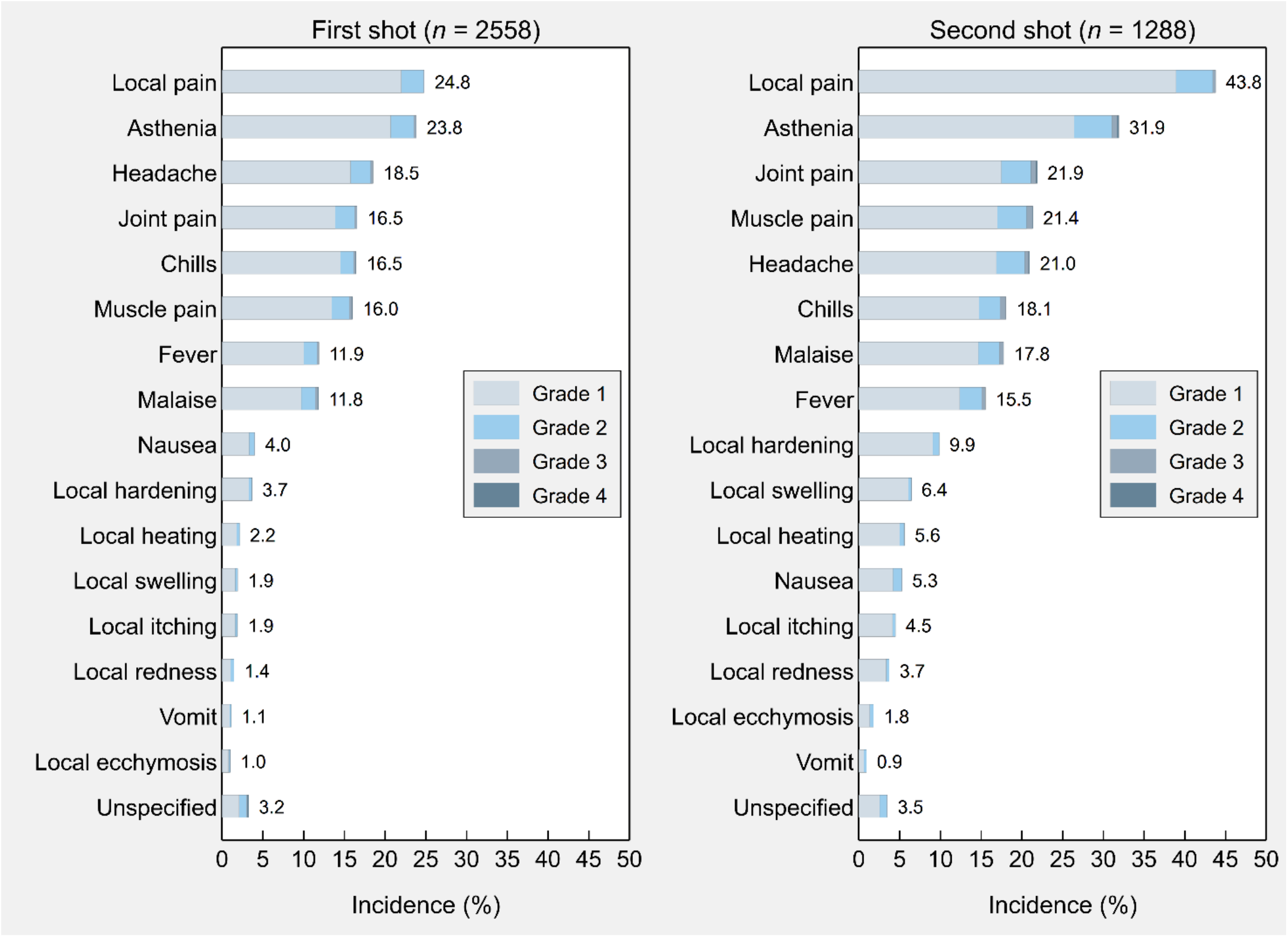
Incidence of specific adverse events, both local and systemic, following immunisation with the first shot and second shot of Sputnik V - Republic of San Marino (2021).

Most symptoms appeared within 24–48 hours after the injection (85.7%), and less frequently after a few minutes (4.5%), 3-5 days (4.2%) and 6–7 days (1.4%); 4.3% of the patients who suffered from AEFI did not provide information about the timing of onset of symptoms.

Overall, we found that the incidence of participants reporting no AEFI progressively increased from 10.4% in the 18-39 age group to 63.2% in the 80-89 age group (Figure 2). AEFI were treated with drugs by 29.2% of the patients (4.1% of missing values).

The 3.4% of the participants referred to have taken painkillers and/or antiinflammatory drugs the day of the vaccination, before receiving it (Table 2).

### Second dose AEFI

About half of the 2,558 participants who answered the Q1 questionnaire, 50.4% (n=1288) filled the Q2 questionnaire after the second dose of the vaccine (Table 3).

**Table 3.** Sociodemographic and clinical characteristics of the individuals who received one dose versus two doses of Sputnik V - Republic of San Marino (2021). Values are counts (percentages) or mean ± standard deviation.

| Characteristic | Only first<br>shot<br>(*n* = 1270) | First and<br>second shot<br>(*n* = 1288) |
| --- | --- | --- |
| Female | 720 (56.7) | 704 (54.7) |
| Age, y | 62.5 ± 13.4 | 68.7 ± 14.5 |
| Age group, y |  |  |
| 19–39 | 106 (8.3) | 77 (6.0) |
| 40–49 | 100 (7.9) | 77 (6.0) |
| 50–59 | 139 (10.9) | 113 (8.8) |
| 60–69 | 576 (45.4) | 210 (16.3) |
| 70–79 | 260 (20.5) | 539 (41.8) |
| 80–89 | 89 (7.0) | 272 (21.1) |
| Underlying medical conditions | 876 (69.0) | 1044 (81.1) |
| Allergies | 318 (25.0) | 309 (24.0) |
| Ongoing drug therapies | 853 (67.2) | 994 (77.2) |
| Infection with SARS-CoV-2 before<br>first vaccination | 52 (4.1) | 41 (3.2) |

Among these, both local and systemic reactions were reported in 31.9% of cases; besides, 18.5% reported systemic reactions, and 16.1% reported local symptoms only. Amongst local reactions, the most reported ones were pain (43.8%), nodules (9.9%), swelling (6.4%), warmth (5.6%). Frequent systemic reported symptoms were asthenia (31.9%), joint pain (21.9%), muscle pain (21.4%), headache (21.0%), chills (18.1%), malaise (17.8%), and fever (15.5%) (Figure 3, 4). For other AEFI, not included in the provided list, see the Table S2.

**Figure 4.**
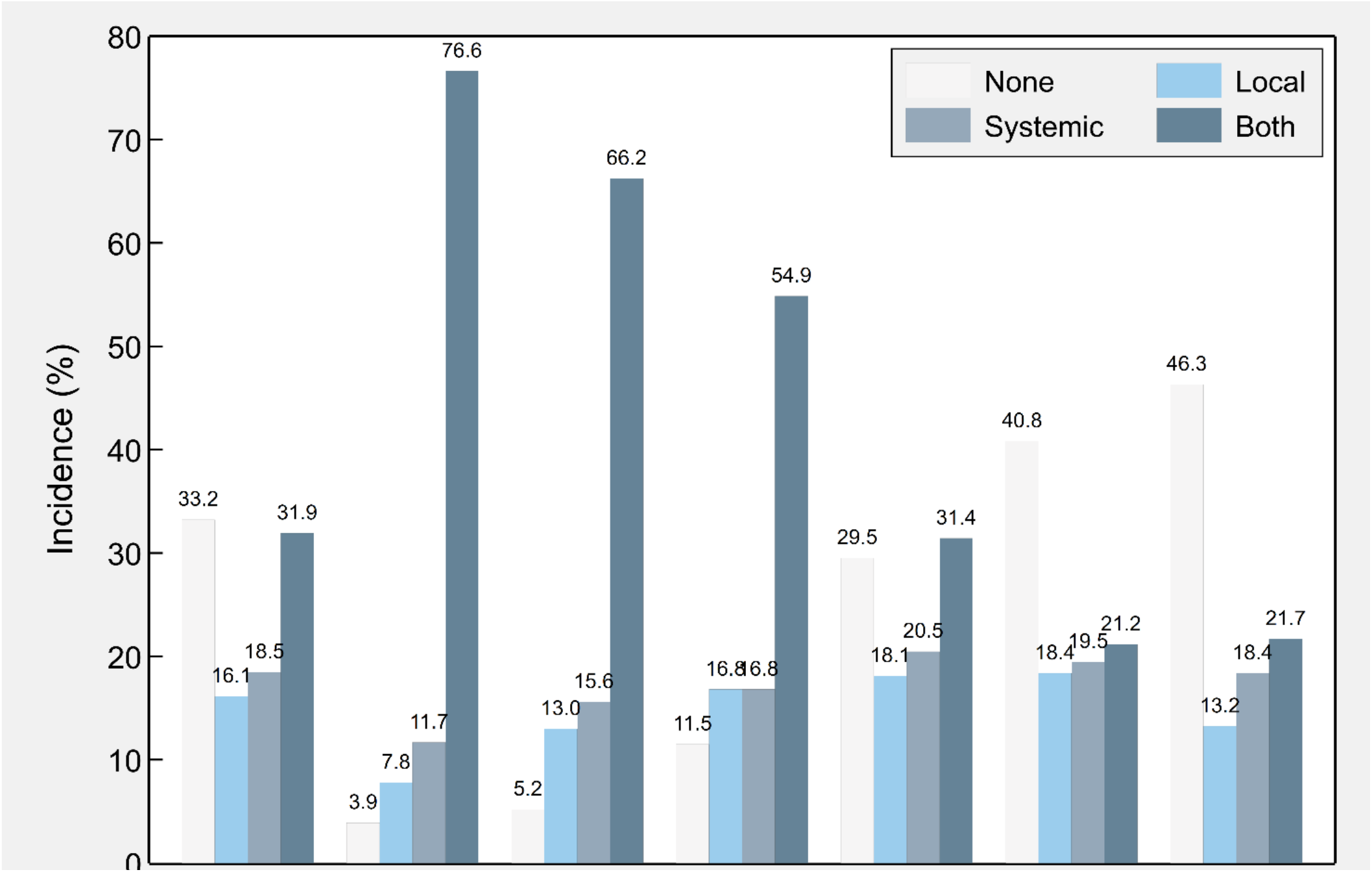
Incidence of local and systemic adverse events following immunisation with the second shot of Sputnik V (n = 1288), overall and by age group - Republic of San Marino (2021). *Notes:* Percentages of each age group do not always add up to 100 because unspecified symptoms were not included in the chart (all: 0.2%; 19–39 y: 0.0%; 40–49 y: 0.0%; 50–59 y: 0.0%; 60–69 y: 0.5%; 70–79 y:0.2%; 80–89 y: 0.4%).

Most symptoms appeared within 24–48 hours after the injection (87.2%), and less frequently after a few minutes (4.3%), 3-5 days (3.4%) and 6–7 days (0.6%); 4.5% of the patients who suffered from AEFI did not provide information about the timing of onset of symptoms.

The incidence of participants reporting no AEFI progressively increased from 3.9% in the 18-39 age group to 46.3% in the 80-89 age group (Figure 4). AEFI were treated with drugs by 27.3% of the patients.

In general, 44 individuals (3.4%) reported having taken painkillers or anti-inflammatory drugs before receiving the second dose of the vaccine (data not shown).

In Figure 5, differences between the two shots of Sputnik V (Gam-COVID-Vac) in the post-injection incidence of local and systemic adverse reactions are shown. In particular, data for over 60 years were statistically significant (p<0.001). The highest incidence (81.8%) of AEFI after the second dose having reported AEFI after the first dose occurred in the age group 60-69, while for those not having reported any AEFI after the first dose we found a 18.2% incidence of AEFI after the second dose in the same age group.

**Figure 5.**
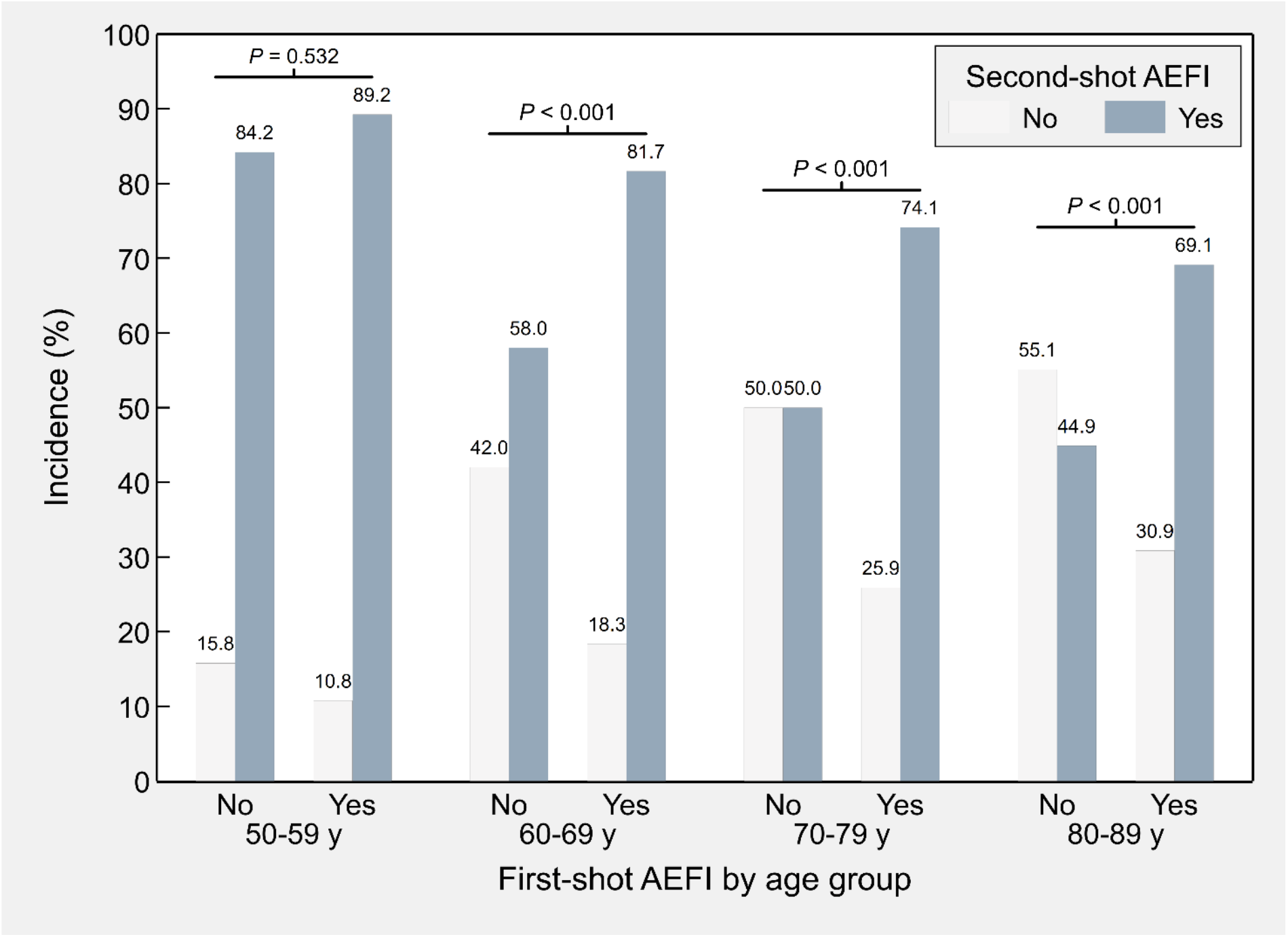
Incidence of specific adverse events, either local or systemic, following immunisation (AEFI) with the second shot of Sputnik V according to the presence or absence of (AEFI) after the first shot, by age group - Republic of San Marino (2021). *Notes: P* values indicate whether the conditional distributions of second-shot AEFI are significantly different according to the presence/absence of first-shot AEFI. Ages <50 years were discarded due to small sample sizes (*n* = 1134).

### AEFI grading

After the first dose of the vaccine, participants reporting at most grade 1 symptoms were 43.5% (n=1,112), at most grade 2 were 8.7% (n=223), at most grade 3 were 0.8% (n=20), and at most grade 4 0.3% (n=8). Grade 4 was reported in 0.1% of the recipients for each chills, headache, and local symptoms. After the second dose of the vaccine, participants reporting at most grade 1 symptoms were 53.8% (n=693), at most grade 2 were 11.3% (n=146), at most grade 3 were 1.3% (n=17), and at most grade 4 0.3% (n=4). Symptoms reported as grade 4 were asthenia (0.2%), headache (0.2%), joint pain (0.2%), chills (0.1%), muscle pain (0.1%), and local symptoms (0.1%) (Table S3, S4).

Amongst over 60 year old vaccine recipients, after the first dose 1.3% (n=17) has reported at most grade 3 AEFI and 0.3% (n=4) at most grade 4 AEFI; for what concerns the second dose, the severity of AEFI was reported by 0.4% (n=4) by 0.3% (n=3) for at most grade 3 and 4 respectively. (Table S3, S4).

## Discussion

The ongoing ROCCA study on Sputnik V (Gam-COVID-Vac) safety is an opportunity to highlight potential AEFI in a real world context of active surveillance, thus mirroring actual incidence. The interim analysis confirmed a good tolerability profile in the over 60 years age group after both doses (rAd26-S and rAd5-S).

Our results align with the findings of phase 1 and 2 previously conducted studies [12], then confirmed by phase 3 of the vaccine trial [9], in terms of overall safety and tolerability. No serious adverse events nor deaths were reported.

Nearly all reported AEFI were mild or moderate and/or lasted less than 2 days, and in more than two thirds of cases no need for any medication was reported. The incidence of participants who reported no AEFI grew consistently along with the progression of age groups, after both doses.

The vast majority of AEFI appeared within 2 days from the inoculation. This applies both to the first and second dose of the vaccine.

Local pain was the most recurrent AEFI both after the first and second dose, followed by asthenia, and headache for the first dose, vs joint pain for the second dose. In particular, the rate of local symptoms is substantially lower than the ones observed in other approved anti-COVID vaccine trials for what concern age groups similar to our sample (>60 years) [13,14]. The majority of AEFI was described as mild (grade 1 and 2), whereas only a few participants reported severe (grade 3 and 4) AEFI. For participants over 60 we can confidently affirm that Sputnik V (Gam-COVID-Vac) showed no safety concerns, having displayed a very limited number of grade 3 and 4 AEFI both after the first and the second dose.

Overall, systemic events had a similar incidence if compared to available data regarding other vaccines used worldwide [13,14,15,16]. Amongst these, asthenia was reported less frequently than for available mRNA vaccines, for both first and second dose, whereas fever and chills showed a higher frequency, especially for the first dose [13,14].

Participants experiencing both local and systemic AEFI increased in the second dose when compared to the first one; when referring to those experiencing only local AEFI, the same increase was especially prominent in older age groups.

Our findings allowed us to point out how, in the population over 60, having reported AEFI after the first dose could be a predictor of AEFI recurrence after the second dose. Amongst those having had an AEFI after the first dose, the probability of AEFI after the second dose decreased with the increasing age. About half of the participants experienced AEFI after the second dose not having had one after the first dose.

The ROCCA study protocol presents several limitations. Being the one we are presenting an interim analysis and considering that the vaccination campaign is still ongoing, the sample of the study is limited. In particular, this compromises the possibility of fully detecting rare AEFI, especially for the subgroups of the population presenting specific comorbidities (eg. diabetes, neurological diseases). Moreover, the youngest age groups are inadequately represented in our sample, as the vaccination campaign is still in the early phases which prioritize the elderly, among others. We plan on resolving said issue as the campaign progresses towards the younger population as scheduled. Finally, our follow-ups still cover a short timeframe and might not be sufficiently distanced from the administration of the vaccine to reveal any delayed AEFI.

The ROCCA vaccine-vigilance study represents the first attempt at investigating AEFI of the Sputnik V vaccine, excluding its phase 1, 2 and 3 trials. As stated earlier, follow-ups to at least 3 months after the administration of the first dose and the inclusion in the vaccination campaign of other age groups will allow us to have a representative sample of the whole population of the Republic of San Marino and to describe any other atypical/rare AEFI. Further studies on the efficacy and effectiveness are clearly needed to complete the overall picture of this vaccine.

The vaccination campaign in the Republic of San Marino began later than in other countries, but it is quickly closing the gaps thanks to the adoption of the Sputnik V vaccine on a mass scale. The demonstration of its safety is another step on the long journey to finally overcome the pandemic and the ROCCA study will keep monitoring it until the end of the campaign.

**BOX 1: What is already known on this topic**

- In December 2020 mass distribution of the recently developed Sputnik V vaccine (Gam-COVID-Vac) started, with the vaccine currently being distributed in 61 countries.
- Phase 1, 2 and 3 trials showed a high efficacy and safety profile, though these data need post-marketing confirmation through dedicated studies.

**BOX 2: What this study adds**

- The interim safety results of the rAd26 and rAd5 vector-based COVID-19 vaccine Gam-COVID-Vac first describe the safe profile in those aged over 60 years after both doses.
- Our results align with the findings of phase 3 of the vaccine trial, in terms of overall safety, in a real world context of active surveillance. The majority of AEFIs reported in our study were mild and moderate, suggesting a high tolerability.

## Data Availability

All data will be available at request sending an e-mail to

## Contributorship statement

All the authors have contributed equally to the conceptualization and design of the manuscript. MM, GS, ZDV, AS, GLF, DG were responsible for drafting the manuscript. JL and MF conducted all data analyses. MM, GS, ZDV, AS, GLF were integral to the design and development of the vaccine safety surveillance questionnaire. MM, GS, ZDV, AS, EB, RM, GLF were involved in the data collection process. All authors made substantial contributions to the interpretation of data for the work and revised the manuscript critically for important intellectual content. All authors had final approval of the version to be published and agreed to be accountable for all aspects of the work in ensuring that questions related to the accuracy or integrity of any part of the work are appropriately investigated and resolved. The corresponding author attests that all listed authors meet authorship criteria and that no others meeting the criteria have been omitted

## Patient consent for publication

Patient consent for publication of this article is not required.

## Transparency Statement

The lead author affirms that this manuscript is an honest, accurate, and transparent account of the study being reported; that no important aspects of the study have been omitted; and that any discrepancies from the study as planned (and, if relevant, registered) have been explained.

## Ethics approval

The study protocol has been reviewed and approved by the Ethics Committee for Research and Experimentation of the Republic of San Marino with the approval number 30/CERS/2021 of 17th of March 2021.

## Data availability statement

The dataset generated and analyzed during the current study can be made available by the corresponding author, AS, on reasonable request.

## Open access

This is an open access article distributed in accordance with the Creative Commons Attribution Non Commercial (CC BY-NC 4.0) license, which permits others to distribute, remix, adapt, build upon this work non-commercially, and license their derivative works on different terms, provided the original work is properly cited and the use is non-commercial.

See: http://creativecommons.org/licenses/by-nc/4.0

## Footnotes

1 **ROCCA**: **R**SM **O**bservatory for **C**OVID vaccination **C**ampaign monitoring **A**dverse events

