## Supplementary Material; Figure S1; Table S1; Table S2; Table S3, S4 for "ROCCA study protocol and interim analysis on safety of Sputnik V vaccine (Gam-COVID-Vac) in the Republic of San Marino: an observational study using active surveillance"

**This appendix has been provided by the authors to give readers additional information about their work.**

**Supplementary Material**

Sommario

### **List of investigators**

| Dr. Alessandra Bruschi | SSI General Director | San Marino |
| --- | --- | --- |
| Dr. Sergio Rabini | SSI Chief Medical Officer | San Marino |
| Dr. Francesca Masi | SSI Prevention Department Director | San Marino |
| Dr. Micaela Santini | SSI Primary Care Director | San Marino |
| Dr. Ivonne Zoffoli | State Hospital Chief Medical Officer | San Marino |
| Dr. Agostino Ceccarini | SSI Primary Care and Territorial Health Director | San Marino |
| Dr. Stefania Stefanelli | Public Relations Office | San Marino |
| Dr. Loredana Stefanelli | Unit for Internal Medicine Director | San Marino |
| Dr. Tatiana Mancini | Unit for Endocrine-metabolic Diseases Director | San Marino |
| Dr. Massimo Arlotti | SSI Consulting Infectious Disease Specialist | San Marino |

### **Figure S1. *Questionnaires***

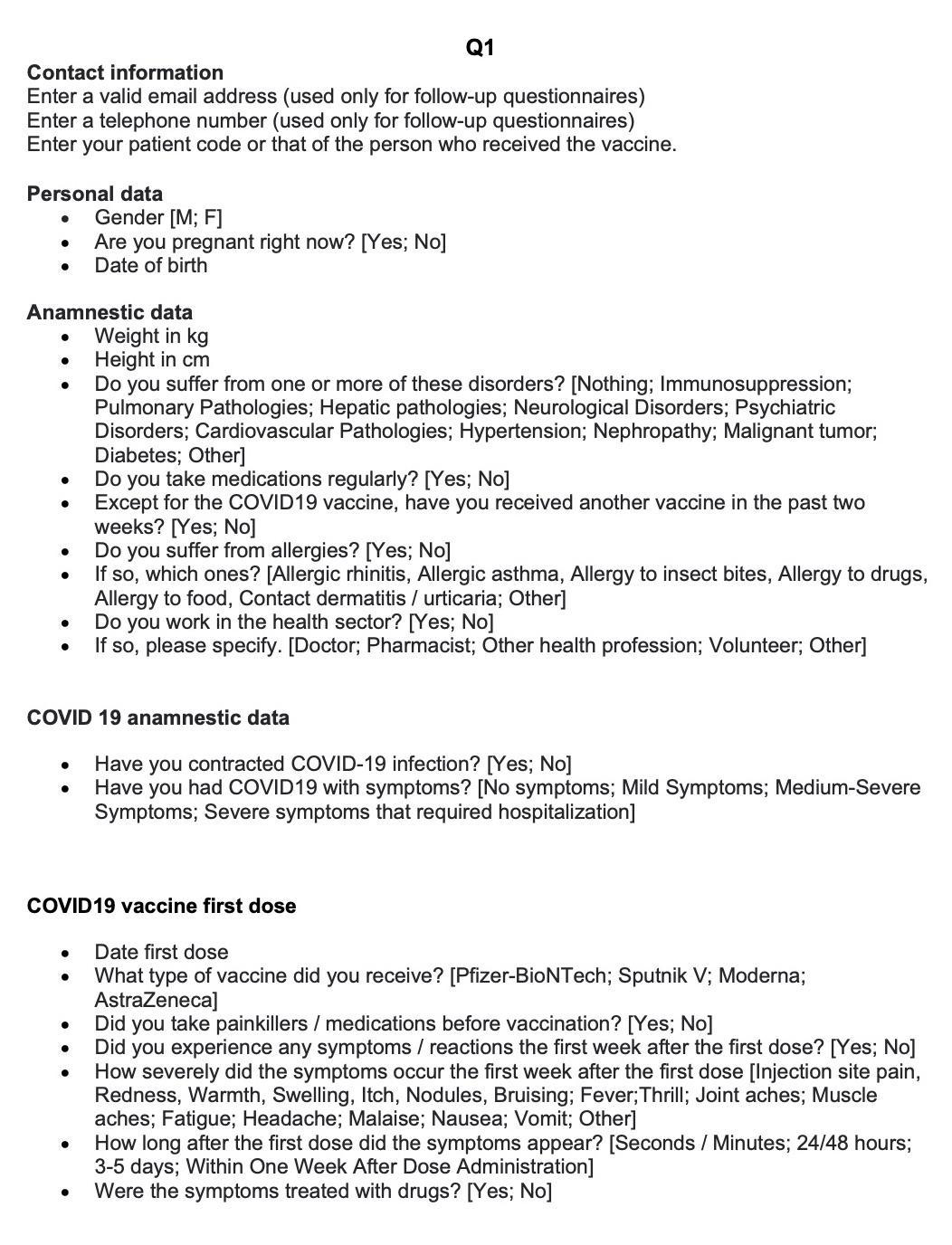

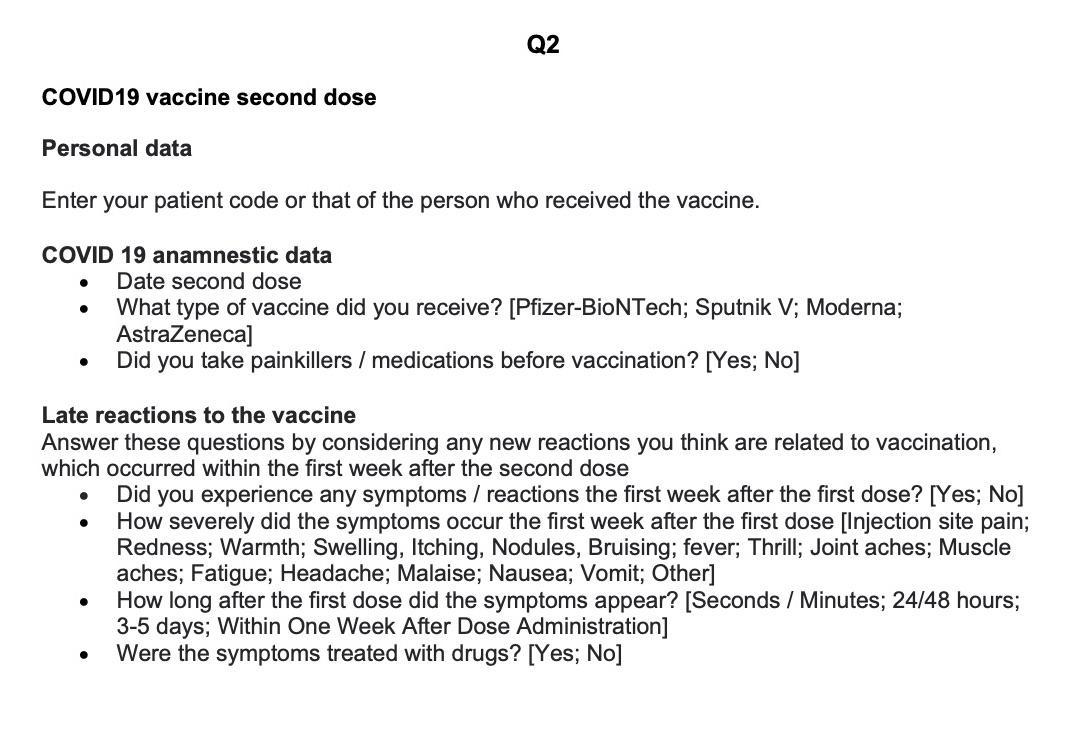

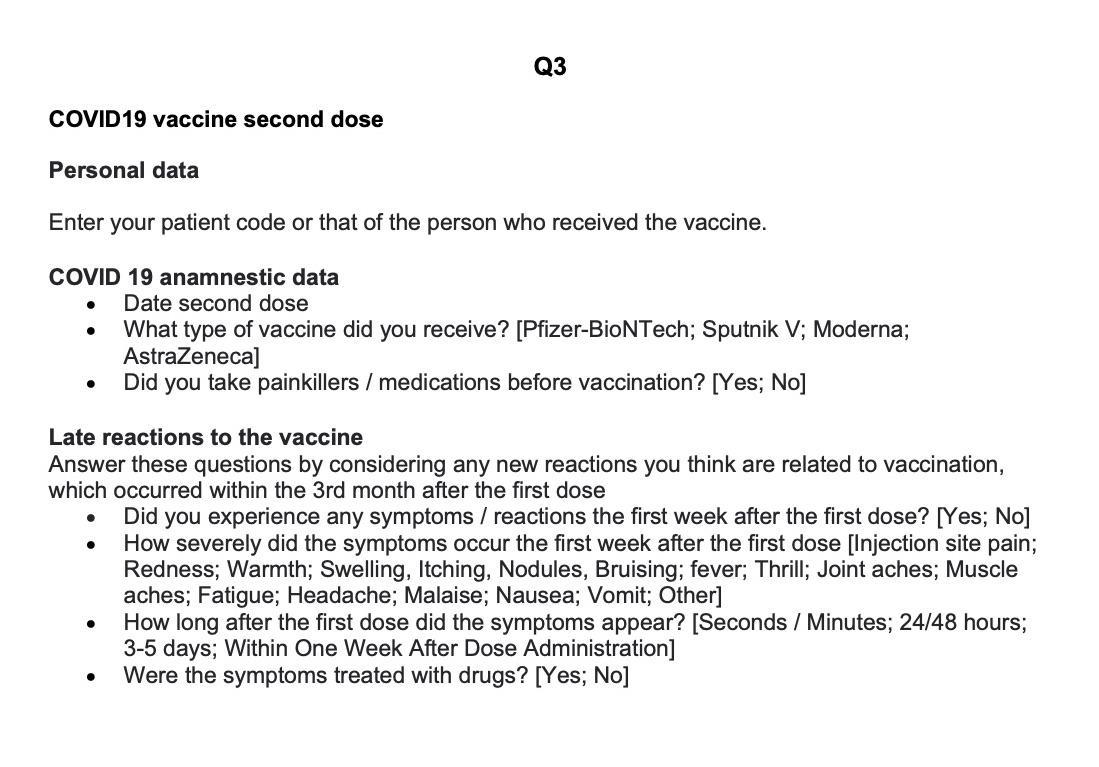

### **Table S1. *First dose atypical AEFI***

| M/F | Time after vaccination | Description |
| --- | --- | --- |
| M | 7 days | Migrant, generalised and progressive itching |
| F | 30 mins | Dyspnea, arterial hypertensive crisis, severe malaise |
| F | 7 days | Dyspnea, low level of blood oxygenation |
| F | 15 mins | Exacerbation of her multiple sclerosis neuropathic pain |
| F | 30 mins | Epiglottitis occlusion and choking sensation |
| M | 24 hours | Several episodes of tachycardia |
| F | respectively 24 hours and 3 days | Neuropathic pain and dermatological rash in the areas of the dermatomes where she suffered from Herpes Zoster years before |
| M | 4 hours | Altered taste feelings |
| F | 3 days | Arterial Hypertensive crisis with admission to the ER |
| M | 12 hours | Dyspnea |
| F | 12 hours | Exacerbation of her usual lumbar neurological pain |
| M | 5 days | Diffuse ortycharioid reaction |
| M | 24 hours | Submental swollen lymph nodes |
| M | 6 days | Diffuse ortycharioid reaction |
| F | 2 hours | Diffuse neuropathic pain |
| F | 12 hours | Arterial Hypertensive crisis |
| F | 1 hours | Tongue edema |
| F | 1 hour | Exacerbation of her usual gastritis algia |

### **Table S2. *Second dose atypical AEFI***

| M/F | Time after vaccination | Description |
| --- | --- | --- |
| F | 48 hours | Hematuria, dyspnea, syncope, abdominal pain, admission to the ER |
| F | 48 hours | Painful and swollen axillary lymph nodes |
| F | 48 hours | Progressing and worsening vertigo |
| F | 48 hours | Diffuse neuropathic pain |
| F | 15 minutes | Burning feeling in the tongue and perioral area |
| F | 36 hours | Arterial hypertensive crisis with cephalea and acouphene |
| F | 3 days | Ortycharioid reaction diffuse on the upper limb not involved in the vaccination |
| F | 12 hours | Vertigo |
| F | 24 hours | Diffuse neuropathic pain |
| F | 12 hours | Insomnia |

### **Table S3. *Incidence of specific adverse events, both local and systemic, following immunisation with the first shot (n = 2558) and the second shot (n = 1288) of Sputnik V, overall and in individuals aged 60–89 years - Republic of San Marino (2021)***

| Adverse event following immunisation | First shot | | | | | | Second shot | | | | | |
| --- | --- | --- | --- | --- | --- | --- | --- | --- | --- | --- | --- | --- |
|  | Rank | Any | Grade | Grade | Grade | Grade | Rank | Any | Grade | Grade | Grade | Grade |
|  | (#) | grade | 1 | 2 | 3 | 4 | (#) | grade | 1 | 2 | 3 | 4 |
| All ages (19–89 y) |  |  |  |  |  |  |  |  |  |  |  |  |
| Local symptoms^*^ | 1 | 26.6% | 23.4% | 3.0% | 0.1% | 0.1% | 1 | 48.1% | 42.5% | 5.1% | 0.3% | 0.1% |
| Asthenia | 2 | 23.8% | 20.7% | 2.8% | 0.3% | 0.0% | 2 | 31.9% | 26.4% | 4.7% | 0.6% | 0.2% |
| Headache | 3 | 18.5% | 15.8% | 2.5% | 0.2% | 0.1% | 5 | 21.0% | 16.8% | 3.5% | 0.5% | 0.2% |
| Joint pain | 4 | 16.5% | 13.9% | 2.3% | 0.3% | 0.0% | 3 | 21.9% | 17.5% | 3.6% | 0.6% | 0.2% |
| Chills | 5 | 16.5% | 14.5% | 1.6% | 0.3% | 0.1% | 6 | 18.1% | 14.8% | 2.6% | 0.7% | 0.1% |
| Muscle pain | 6 | 16.0% | 13.4% | 2.2% | 0.4% | 0.0% | 4 | 21.4% | 17.0% | 3.6% | 0.7% | 0.1% |
| Fever | 7 | 11.9% | 10.0% | 1.6% | 0.2% | 0.0% | 8 | 15.5% | 12.3% | 2.7% | 0.5% | 0.0% |
| Malaise | 8 | 11.8% | 9.7% | 1.7% | 0.4% | 0.0% | 7 | 17.8% | 14.7% | 2.6% | 0.5% | 0.0% |
| Nausea | 9 | 4.0% | 3.3% | 0.6% | 0.1% | 0.0% | 9 | 5.3% | 4.2% | 1.0% | 0.1% | 0.0% |
| Vomit | 10 | 1.1% | 0.9% | 0.2% | 0.0% | 0.0% | 10 | 0.9% | 0.6% | 0.3% | 0.0% | 0.0% |
| Unspecified | · | 3.2% | 2.1% | 0.9% | 0.1% | 0.2% | · | 3.5% | 2.6% | 0.9% | 0.1% | 0.0% |
| Age 60–89 y |  |  |  |  |  |  |  |  |  |  |  |  |
| Local symptoms^*^ | 1 | 18.1% | 16.3% | 1.7% | 0.0% | 0.1% | 1 | 40.4% | 36.9% | 3.2% | 0.1% | 0.1% |
| Asthenia | 2 | 15.2% | 13.9% | 1.3% | 0.1% | 0.0% | 2 | 23.7% | 21.1% | 2.4% | 0.0% | 0.2% |
| Headache | 3 | 12.0% | 10.7% | 1.2% | 0.1% | 0.0% | 5 | 13.0% | 11.5% | 1.3% | 0.1% | 0.2% |
| Joint pain | 4 | 10.0% | 8.9% | 1.1% | 0.0% | 0.0% | 3 | 14.9% | 12.7% | 1.8% | 0.2% | 0.2% |
| Chills | 5 | 9.4% | 9.0% | 0.3% | 0.0% | 0.0% | 6 | 12.5% | 11.6% | 0.6% | 0.3% | 0.1% |
| Muscle pain | 6 | 9.1% | 8.3% | 0.8% | 0.0% | 0.0% | 4 | 13.9% | 11.8% | 1.8% | 0.3% | 0.1% |
| Fever | 7 | 6.1% | 5.8% | 0.4% | 0.0% | 0.0% | 7 | 9.7% | 8.6% | 0.9% | 0.2% | 0.0% |
| Malaise | 8 | 5.7% | 5.2% | 0.4% | 0.1% | 0.0% | 8 | 9.3% | 8.6% | 0.6% | 0.1% | 0.0% |
| Nausea | 9 | 2.5% | 2.2% | 0.3% | 0.1% | 0.0% | 9 | 3.1% | 2.9% | 0.2% | 0.0% | 0.0% |
| Vomit | 10 | 0.8% | 0.7% | 0.1% | 0.0% | 0.0% | 10 | 0.6% | 0.5% | 0.1% | 0.0% | 0.0% |
| Unspecified | · | 2.3% | 1.4% | 0.6% | 0.2% | 0.2% | · | 3.3% | 2.5% | 0.8% | 0.0% | 0.0% |

* Patients with multiple symptoms were assigned the highest grade reported.

**Table S4. *Percentage distribution of symptom severity (1 to 4) of adverse events following immunisation with the first and second shot of Sputnik V, overall and in individuals aged 60–89 years - Republic of San Marino (2021).***

| Adverse event following immunisation | First shot | | | | | | Second shot | | | | | |
| --- | --- | --- | --- | --- | --- | --- | --- | --- | --- | --- | --- | --- |
|  | Rank | *n* | Symptom severity % distribution | | | | Rank | *n* | Symptom severity % distribution | | | |
|  | (#) |  | 1 | 2 | 3 | 4 | (#) |  | 1 | 2 | 3 | 4 |
| All ages (18–89 y) |  |  |  |  |  |  |  |  |  |  |  |  |
| Local symptoms^*^ | 1 | 681 | 88.0% | 11.5% | 0.3% | 0.3% | 1 | 619 | 88.5% | 10.7% | 0.6% | 0.2% |
| Asthenia | 2 | 610 | 86.7% | 11.8% | 1.3% | 0.2% | 2 | 411 | 82.7% | 14.6% | 1.9% | 0.7% |
| Headache | 3 | 474 | 85.0% | 13.3% | 1.3% | 0.4% | 5 | 270 | 80.4% | 16.7% | 2.2% | 0.7% |
| Joint pain | 4 | 423 | 83.9% | 14.2% | 1.9% | 0.0% | 3 | 282 | 79.8% | 16.7% | 2.8% | 0.7% |
| Chills | 5 | 421 | 88.4% | 9.5% | 1.7% | 0.5% | 6 | 233 | 81.5% | 14.2% | 3.9% | 0.4% |
| Muscle pain | 6 | 409 | 84.1% | 13.4% | 2.2% | 0.2% | 4 | 275 | 79.6% | 16.7% | 3.3% | 0.4% |
| Fever | 7 | 304 | 84.5% | 13.2% | 2.0% | 0.3% | 8 | 200 | 79.5% | 17.5% | 3.0% | 0.0% |
| Malaise | 8 | 302 | 82.5% | 14.2% | 3.0% | 0.3% | 7 | 229 | 82.5% | 14.4% | 3.1% | 0.0% |
| Nausea | 9 | 103 | 82.5% | 15.5% | 1.9% | 0.0% | 9 | 68 | 79.4% | 19.1% | 1.5% | 0.0% |
| Vomit | 10 | 28 | 82.1% | 17.9% | 0.0% | 0.0% | 10 | 12 | 66.7% | 33.3% | 0.0% | 0.0% |
| Unspecified | · | 83 | 63.9% | 27.7% | 3.6% | 4.8% | · | 45 | 73.3% | 24.4% | 2.2% | 0.0% |
| Age 60–89 y |  |  |  |  |  |  |  |  |  |  |  |  |
| Local symptoms^*^ | 1 | 353 | 90.1% | 9.6% | 0.0% | 0.3% | 1 | 412 | 91.5% | 8.0% | 0.2% | 0.2% |
| Asthenia | 2 | 296 | 91.2% | 8.4% | 0.3% | 0.0% | 2 | 242 | 88.8% | 10.3% | 0.0% | 0.8% |
| Headache | 3 | 233 | 89.3% | 9.9% | 0.9% | 0.0% | 5 | 133 | 88.0% | 9.8% | 0.8% | 1.5% |
| Joint pain | 4 | 195 | 89.2% | 10.8% | 0.0% | 0.0% | 3 | 152 | 85.5% | 11.8% | 1.3% | 1.3% |
| Chills | 5 | 182 | 96.7% | 3.3% | 0.0% | 0.0% | 6 | 128 | 92.2% | 4.7% | 2.3% | 0.8% |
| Muscle pain | 6 | 177 | 91.0% | 9.0% | 0.0% | 0.0% | 4 | 142 | 84.5% | 12.7% | 2.1% | 0.7% |
| Fever | 7 | 119 | 94.1% | 5.9% | 0.0% | 0.0% | 7 | 99 | 88.9% | 9.1% | 2.0% | 0.0% |
| Malaise | 8 | 111 | 91.9% | 6.3% | 1.8% | 0.0% | 8 | 95 | 92.6% | 6.3% | 1.1% | 0.0% |
| Nausea | 9 | 48 | 87.5% | 10.4% | 2.1% | 0.0% | 9 | 32 | 93.8% | 6.3% | 0.0% | 0.0% |
| Vomit | 10 | 15 | 93.3% | 6.7% | 0.0% | 0.0% | 10 | 6 | 83.3% | 16.7% | 0.0% | 0.0% |
| Unspecified | · | 45 | 60.0% | 26.7% | 6.7% | 6.7% | · | 34 | 76.5% | 23.5% | 0.0% | 0.0% |

* Patients with multiple symptoms were assigned the highest grade reported.
